# Association between relative income and mental health in adults during the COVID-19 pandemic in Korea: Insights from a community health survey

**DOI:** 10.1101/2023.07.14.23292685

**Authors:** Min Hui Moon, Min Hyeok Choi

**Affiliations:** Department of Preventive and Occupational & Environmental Medicine, Medical College, Pusan National University, Yangsan, Republic of Korea; Office of Public Healthcare Service, Pusan National University Yangsan Hospital, Yangsan, Republic of Korea

## Abstract

People of lower socioeconomic status are much more likely to be vulnerable to COVID-19. This study aimed to compare the associations between mental health according to relative national and community income levels during the COVID-19 pandemic. Mental health inequalities according to income level during the COVID-19 pandemic were assessed using the Korea Community Health Survey before (2019) and after the COVID-19 pandemic (2021). Univariate analyses were used to calculate the perceived stress and depression rates according to the risk factor categories. A multivariate logistic regression analysis was performed to identify the association between two types of income levels (Korean or community) and perceived stress and depression. In addition, we investigated the effect of relative income levels by subgroup (gender and region) on perceived stress and the experience of depression. During COVID-19, although depression crude rates increased (from 6.24% to 7.2%), perceived stress crude rates remained similar. In addition, as for mental health inequality according to community income level, even after adjusting for each independent variable, perceived stress [Odds Ratio (OR): 1.31, 95% Confidence Interval (CI):1.31–1.32] and experience of depression (OR: 1.63, 95% CI: 1.62–1.63) increased as the income level decreased. The effect of relative income level on perceived stress rate was found to be more pronounced in urban areas than in rural areas in terms of community income levels. Contrarily, the effect of relative income level on the depression rate was found to be weaker. Our findings demonstrated that mental health inequalities based on income level were more likely to occur during the COVID-19 pandemic and that disparities in community income levels may better reflect mental health inequalities.

## Introduction

The coronavirus disease (COVID-19) pandemic has significantly impacted mental health worldwide, including stress, anxiety, and depression worldwide [1,2]. According to a 2021 OECD report (Hewlett, E. et al.), the prevalence of depression in OECD countries has approximately doubled since the COVID-19 pandemic, with the prevalence being the highest in Korea at 36.8% among OECD countries [3]. During the COVID-19 pandemic, social distancing and containment measures had a direct impact on mental health [4–6]. Several previous studies have reported that large-scale infectious diseases can cause emotional confusion and difficulties such as depression and anxiety [7–12]. Cao et al. [7] and Shevlin et al. [11] explored the effects of the COVID-19 pandemic and social isolation on mental health, and Lee et al. [9] investigated factors related to fear of the COVID-19 infection and its psychological and social impact. Furthermore, studies have shown that social isolation due to COVID-19 and fear and awareness of the infection increases symptoms of depression and anxiety [4].

Mental health related to stress and depression is affected by socioeconomic risk factors such as education level, occupation, and income level. According to Patel et al.’s [13] study on the effect of income level on mental health inequality among representative socioeconomic factors, 33 surveys conducted in 20 countries reported that the lower the income level, the higher the risk of depression. Hong et al. [14] found that inequality based on income level is more pronounced in mental health than in physical health, doubling the size of inequality over 10 years. Several other studies have consistently shown that people with lower socioeconomic status are more vulnerable to mental health problems [13–18]. Several studies have identified mental health inequality according to income level at the national level. Meanwhile, Song A and Kim W [15] studied income inequality at the national as well as community levels. As income inequality at the community level has a significant impact on social capital and access to healthcare infrastructure, measuring health gaps is more useful than income inequality at the national level [15,16].

The COVID-19 pandemic has exacerbated health inequalities among populations with low socioeconomic status. For example, the lower the education level, income level, or unstable employment status, the higher the risk of COVID-19 infection, and the higher the critical severity and mortality rate [19–25]. Hall et al. [19] examined the impact of income inequality on daily life and mental health during the COVID-19 pandemic. The results showed that the low-income population had difficulty purchasing food and daily necessities, and their health status deteriorated because of a lack of time and resources for proper healthcare. Owing to this influence, the low-income group showed unstable mental health conditions, such as stress and depression, compared to the high-income group.

Previous studies have confirmed that a) mental health issues, such as stress and depression, deteriorated due to the COVID-19 pandemic, and b) there were differences in mental health according to income level [13–15]. These studies were often limited to income inequality at the national level; few studies have analyzed the relationship between income inequality and mental health at the community level, which can reflect the accessibility of healthcare infrastructure in the region.

This study aimed to identify changes in mental health according to income level before (2019) and after (2021) the COVID-19 pandemic. In addition, it was conducted to provide a policy basis for improving mental health inequality by comparing the patterns of health inequality according to income at the national and community levels.

Therefore, the purpose of this study was to:

1) Identify and compare levels of perceived stress and depression according to income level during the COVID-19 pandemic.
2) During the COVID-19 pandemic, we analyzed the relationship between perceived stress and the experience of depression on mental health according to the relative income levels of the total (Korean income level) and local (community income level) population.
3) Investigate the effect of relative income levels by subgroup (gender and region) on perceived stress and experiences of depression.

## Materials and methods

### Data and study population

Data were obtained from the Korea Community Health Survey (KCHS). Since 2008, the Community Health Survey (CHS) has been conducted annually by the Korea Centers for Disease Control and Prevention. The CHS is a large-scale survey in which about 220,000 people nationwide participate from August to October every year and includes questions on chronic disease screening, health behavior, food intake, and socioeconomic status. Survey data were used as official national indicators such as health level, health behavior, food and nutrition intake, and chronic disease prevalence in Korea (Korea Community Health Survey Guidelines, website: http://chs.kdca.go.kr/). This survey was conducted in the form of a 1:1 interview with a surveyor visiting households of adults aged 19 or older residing in 255 cities, counties, and districts in Korea. Research participants were selected through probability proportional sampling and systematic sampling every year. This study selected 449,234 people, excluding those who did not respond, as the final study participants out of 458,341 people who participated in community health surveys in 2019 and 2021.

## Variables

### Dependent variables

The dependent variables included perceived stress and depression. Perceived stress and depression are representative indicators of mental health, and stress plays an important role in predicting depression [26–28]. Several studies have demonstrated that exposure to perceived stress and experiences of depression are associated with poor health outcomes and affect socioeconomic imbalance [29–31]. Low socioeconomic status is associated with a high prevalence of stress and depression; mental health in low-income groups is particularly aggravated by persistent poverty and income inequality [32–34]. Perceived stress was assessed using the question, “How stressful do you feel in your daily life?” with response options of “feel very much,” “feel a lot,” “feel a little bit,” and “hardly feel it.”. For the analysis, those who responded “I feel it very much” and “I feel it a lot” were classified as those who usually feel stress in my daily life, and those who answered “I feel it a little” and “I hardly feel it” was classified as a person who doesn’t. The experience of depression was surveyed using the following question: “Did you feel sadness or despair enough to bother you in your daily life for more than 2 weeks in the last year?” Their answer was recorded as “yes” or “no.” In this study, a participant was defined as one who answered “yes.”

### Independent variables

The health inequality variable, considered a major factor in this study, was income level. The income level is an indicator of socioeconomic status that can be used to directly measure the material resources available to individuals. It is a representative indicator of socioeconomic inequality and is widely used because it implies that income inequality affects health outcomes. Income level is closely related to health; the lower the income level, the higher the rate of unhealthy states [35,36]. The income level used in this study was the equalized income calculated by dividing household income by the square root of the household members. The Korean income level reconstructs the average income level of the entire population into the third quintile, and the community income level is set by dividing it into third quintiles based on the average income level of 255 cities, counties, and districts.

The general factors associated with perceived stress and experience of depression were included as independent variables after reviewing studies that previously reported mental health risk factors [37–39]. Demographic variables included gender (men or women), age group (19–29, 30–64, or ≥65), and area of residence (urban or rural). Social-economic parameters included education level (≤middle school, ≤high school, or college or above), job status (economic activity or non-economic activity), marital status (married or not married) and the basic livelihood condition. Health behavior factors included current smoking (yes or no), high-risk drinking (men: drinking seven standard drinks or more over once a week, women: drinking five standard drinks or more over once a week), and walking practice (walking activity for ≥ 30 min, ≥ five days in the previous week).

### Statistical analysis

Perceived stress and depression rates were calculated by performing univariate analysis according to the dependent variables. Statistically significant differences in perceived stress and experiences of depression were verified by performing the Rao-Scott chi-square test. The association between two types of income levels (Korean and community income levels) and mental health (perceived stress and experience of depression) was analyzed by performing a complex-sample multivariate logistic regression analysis to adjust for other variables. Subgroup analysis was performed based on two types of income levels (Korean and community income levels), gender, and area of residence. Statistical significance was set at a p-value of <0.05. All statistical analyses were performed using SAS 9.4 (SAS Institute, Cary, NC, USA).

### Ethical considerations

This study was approved by the Institutional Review Board of Pusan National University Hospital (IRB No. 04-2022-030). All the participants provided written informed consent for the KCHS. The survey was conducted after sufficiently explaining to the participants that the results would be used for statistical purposes only and that confidentiality was guaranteed. The need for informed consent was waived by the IRB because the data were analyzed anonymously.

## Results

Table 1 shows the general characteristics of the study population. The total number of participants was 449,234, and the number of weighted analysis participants was 84,491,967 (41,590,294 in 2019 and 42,901,673 in 2021). The proportions of men and women were similar (49.66% in 2019 and 49.62% in 2021), and those aged 30–64 years were the most common subgroup population (63.73% in 2019 and 62.75% in 2021). Regarding population distribution by Korean income levels, Q1 (high) had the highest, followed by Q2 and Q3. The same was found for community income levels. The perceived stress rates were 24.78% in 2019 and 2021. The experience of depression rates are 6.25% and 7.2% in 2019 and 2021, respectively.

**Table 1.**
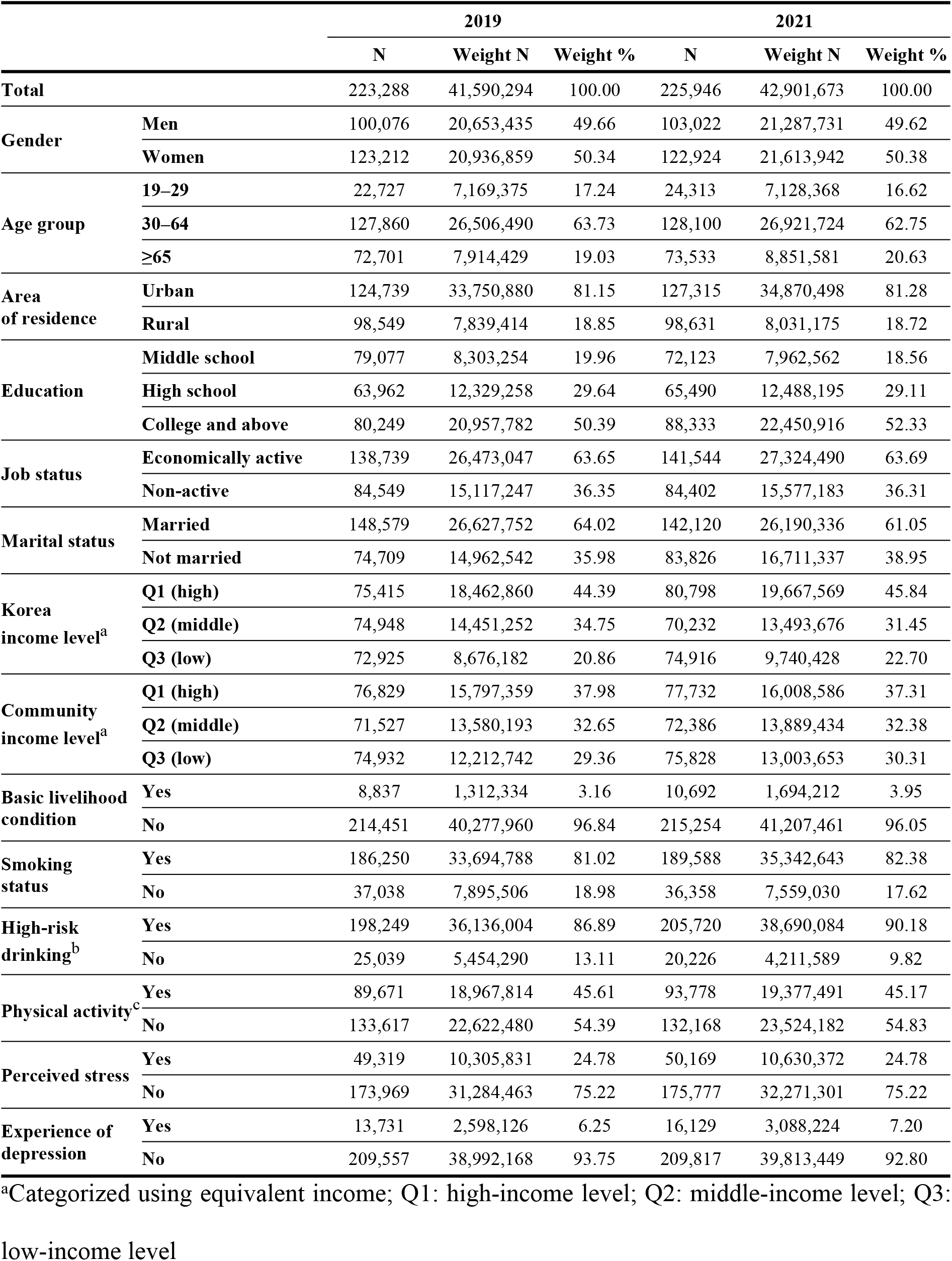
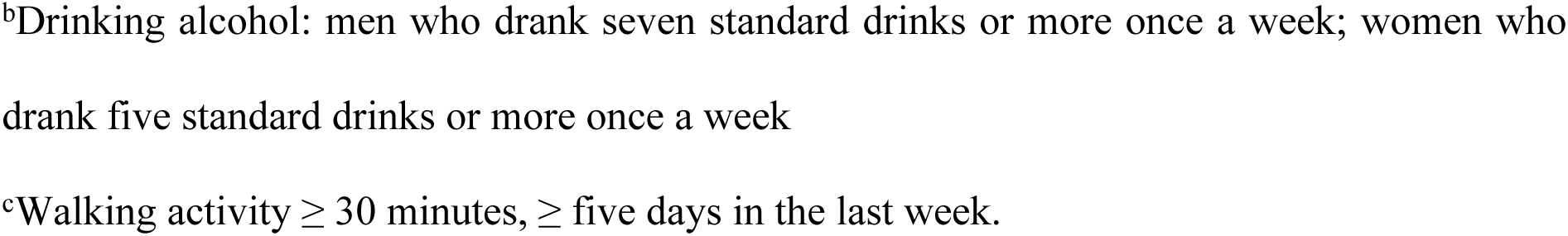
General characteristics of the study population.

|  |  | 2019 |  |  | 2021 |  |  |
| --- | --- | --- | --- | --- | --- | --- | --- |
|  |  | N | Weight N | Weight % | N | Weight N | Weight % |
| **Total** |  | 223,288 | 41,590,294 | 100.00 | 225,946 | 42,901,673 | 100.00 |
| **Gender** | **Men** | 100,076 | 20,653,435 | 49.66 | 103,022 | 21,287,731 | 49.62 |
|  | **Women** | 123,212 | 20,936,859 | 50.34 | 122,924 | 21,613,942 | 50.38 |
| **Age group** | **19–29** | 22,727 | 7,169,375 | 17.24 | 24,313 | 7,128,368 | 16.62 |
|  | **30–64** | 127,860 | 26,506,490 | 63.73 | 128,100 | 26,921,724 | 62.75 |
|  | **≥65** | 72,701 | 7,914,429 | 19.03 | 73,533 | 8,851,581 | 20.63 |
| **Area of residence** | **Urban** | 124,739 | 33,750,880 | 81.15 | 127,315 | 34,870,498 | 81.28 |
|  | **Rural** | 98,549 | 7,839,414 | 18.85 | 98,631 | 8,031,175 | 18.72 |
| **Education** | **Middle school** | 79,077 | 8,303,254 | 19.96 | 72,123 | 7,962,562 | 18.56 |
|  | **High school** | 63,962 | 12,329,258 | 29.64 | 65,490 | 12,488,195 | 29.11 |
|  | **College and above** | 80,249 | 20,957,782 | 50.39 | 88,333 | 22,450,916 | 52.33 |
| **Job status** | **Economically active** | 138,739 | 26,473,047 | 63.65 | 141,544 | 27,324,490 | 63.69 |
|  | **Non-active** | 84,549 | 15,117,247 | 36.35 | 84,402 | 15,577,183 | 36.31 |
| **Marital status** | **Married** | 148,579 | 26,627,752 | 64.02 | 142,120 | 26,190,336 | 61.05 |
|  | **Not married** | 74,709 | 14,962,542 | 35.98 | 83,826 | 16,711,337 | 38.95 |
| **Korea income level<sup>a</sup>** | **Q1 (high)** | 75,415 | 18,462,860 | 44.39 | 80,798 | 19,667,569 | 45.84 |
|  | **Q2 (middle)** | 74,948 | 14,451,252 | 34.75 | 70,232 | 13,493,676 | 31.45 |
|  | **Q3 (low)** | 72,925 | 8,676,182 | 20.86 | 74,916 | 9,740,428 | 22.70 |
| **Community income level<sup>a</sup>** | **Q1 (high)** | 76,829 | 15,797,359 | 37.98 | 77,732 | 16,008,586 | 37.31 |
|  | **Q2 (middle)** | 71,527 | 13,580,193 | 32.65 | 72,386 | 13,889,434 | 32.38 |
|  | **Q3 (low)** | 74,932 | 12,212,742 | 29.36 | 75,828 | 13,003,653 | 30.31 |
| **Basic livelihood condition** | **Yes** | 8,837 | 1,312,334 | 3.16 | 10,692 | 1,694,212 | 3.95 |
|  | **No** | 214,451 | 40,277,960 | 96.84 | 215,254 | 41,207,461 | 96.05 |
| **Smoking status** | **Yes** | 186,250 | 33,694,788 | 81.02 | 189,588 | 35,342,643 | 82.38 |
|  | **No** | 37,038 | 7,895,506 | 18.98 | 36,358 | 7,559,030 | 17.62 |
| **High-risk drinking<sup>b</sup>** | **Yes** | 198,249 | 36,136,004 | 86.89 | 205,720 | 38,690,084 | 90.18 |
|  | **No** | 25,039 | 5,454,290 | 13.11 | 20,226 | 4,211,589 | 9.82 |
| **Physical activity<sup>c</sup>** | **Yes** | 89,671 | 18,967,814 | 45.61 | 93,778 | 19,377,491 | 45.17 |
|  | **No** | 133,617 | 22,622,480 | 54.39 | 132,168 | 23,524,182 | 54.83 |
| **Perceived stress** | **Yes** | 49,319 | 10,305,831 | 24.78 | 50,169 | 10,630,372 | 24.78 |
|  | **No** | 173,969 | 31,284,463 | 75.22 | 175,777 | 32,271,301 | 75.22 |
| **Experience of depression** | **Yes** | 13,731 | 2,598,126 | 6.25 | 16,129 | 3,088,224 | 7.20 |
|  | **No** | 209,557 | 38,992,168 | 93.75 | 209,817 | 39,813,449 | 92.80 |

<sup>a</sup>Categorized using equivalent income; Q1: high-income level; Q2: middle-income level; Q3:
low-income level
<sup>b</sup>Drinking alcohol: men who drank seven standard drinks or more once a week; women who drank five standard drinks or more once a week
<sup>c</sup>Walking activity $\geq 30$ minutes, ≥ five days in the last week.

Table 2 shows perceived stress and experiences of depression according to the factors identified in 2019 and 2021. Perceived stress is significantly higher in women than in men in both 2019 and 2021 (p<0.0001). It was most common in the 30–64 subgroup population, followed by those aged 19–29 and ≥65 years. Perceived stress according to the Korean income level was significantly higher as the relative income level increased, and the community income level was also confirmed by the same result. The number of people living in urban areas was higher than in rural areas. Economic activity was significantly higher than in the non-economic activity group (p<0.0001). Depression was significantly higher in women than in men in both 2019 and 2021 and occurred most commonly in the ≥65 years age group. In 2019 and 2021, the experience of depression rates of lowest Korean income group (Q3) is significantly higher. Community income levels showed the same results (p<0.0001).

**Table 2.** Perceived stress and experience of depression according to factors during the COVID-19 pandemic.

|  |  | Perceived stress |  |  |  | Experience of depression |  |  |  |
| --- | --- | --- | --- | --- | --- | --- | --- | --- | --- |
|  |  | 2019 |  | 2021 |  | 2019 |  | 2021 |  |
|  |  | N | % | N | % | N | % | N | % |
| Gender | Men | 20,789 | 20.77 | 21,384 | 20.76 | 4,350 | 4.35 | 5,315 | 5.16 |
|  | Women | 28,530 | 23.16 | 28,785 | 23.42 | 9,381 | 7.61 | 10,814 | 8.80 |
|  | *p-value*<sup>a</sup> | <.0001 |  | <.0001 |  | <.0001 |  | <.0001 |  |
| Age group | 19–29 | 6,164 | 27.12 | 6,377 | 26.23 | 1,212 | 5.33 | 1,479 | 6.08 |
|  | 30–64 | 31,118 | 24.34 | 32,499 | 25.37 | 7,385 | 5.78 | 8,845 | 6.90 |
| | $\geq 65$ | 12,037 | 16.56 | 11,293 | 15.36 | 5,134 | 7.06 | 5,805 | 7.89 |
|  | *p-value* | <.0001 |  | <.0001 |  | <.0001 |  | <.0001 |  |
| Area of residence | Urban | 29,908 | 23.98 | 30,809 | 24.20 | 8,002 | 6.41 | 9,513 | 7.47 |
|  | Rural | 19,411 | 19.70 | 19,360 | 19.63 | 5,729 | 5.81 | 6,616 | 6.71 |
|  | *p-value* | <.0001 |  | <.0001 |  | <.0001 |  | <.0001 |  |
| Education | Middle school | 14,977 | 18.94 | 12,720 | 17.64 | 6,071 | 7.68 | 6,431 | 8.92 |
|  | High school | 14,005 | 21.90 | 14,807 | 22.61 | 3,859 | 6.03 | 4,613 | 7.04 |
|  | College and above | 20,337 | 25.34 | 22,642 | 25.63 | 3,801 | 4.74 | 5,085 | 5.76 |
|  | *p-value* | <.0001 |  | <.0001 |  | <.0001 |  | <.0001 |  |
| Job status | Economically active | 32,579 | 23.48 | 33,707 | 23.81 | 6,539 | 4.71 | 8,034 | 5.68 |
|  | Non-active | 16,740 | 19.80 | 16,462 | 19.50 | 7,192 | 8.51 | 8,095 | 9.59 |
|  | *p-value* | <.0001 |  | <.0001 |  | <.0001 |  | <.0001 |  |
| Marital status | Married | 31,855 | 21.44 | 30,680 | 21.59 | 7,679 | 5.17 | 8,507 | 5.99 |
|  | Not married | 17,464 | 23.38 | 19,489 | 23.25 | 6,052 | 8.1 | 7,622 | 9.09 |
|  | *p-value* | <.0001 |  | <.0001 |  | <.0001 |  | <.0001 |  |
| Korean income level | Q1 (high) | 17,713 | 23.49 | 19,217 | 23.78 | 3,328 | 4.41 | 4,217 | 5.22 |
|  | Q2 (middle) | 16,244 | 21.67 | 15,456 | 22.01 | 4,071 | 5.43 | 4,381 | 6.24 |
|  | Q3 (low) | 15,362 | 21.07 | 15,496 | 20.68 | 6,332 | 8.68 | 7,531 | 10.05 |
|  | *p-value* | <.0001 |  | <.0001 |  | <.0001 |  | <.0001 |  |
| Community income level | Q1 (high) | 17,237 | 22.44 | 17,670 | 22.73 | 3,313 | 4.31 | 3,964 | 5.10 |
|  | Q2 (middle) | 15,551 | 21.74 | 16,074 | 22.21 | 3,786 | 5.29 | 4,523 | 6.25 |
|  | Q3 (low) | 16,531 | 22.06 | 16,425 | 21.66 | 6,632 | 8.85 | 7,642 | 10.08 |
|  | *p-value* | <.0001 |  | <.0001 |  | <.0001 |  | <.0001 |  |
| Basic livelihood condition | Yes | 2,761 | 31.24 | 3,132 | 29.29 | 1,443 | 16.33 | 1,930 | 18.05 |
|  | No | 46,558 | 21.71 | 47,037 | 21.85 | 12,288 | 5.73 | 14,199 | 6.60 |
|  | *p-value* | <.0001 |  | <.0001 |  | <.0001 |  | <.0001 |  |
| Smoking status | Yes | 10,406 | 28.10 | 10,288 | 28.30 | 2,391 | 6.46 | 2,776 | 7.64 |
|  | No | 38,913 | 20.89 | 39,881 | 21.04 | 11,340 | 6.09 | 13,353 | 7.04 |
|  | *p-value* | <.0001 |  | <.0001 |  | <.0001 |  | <.0001 |  |
| High-risk drinking | Yes | 6,887 | 27.51 | 5,745 | 28.40 | 1,418 | 5.66 | 1,439 | 7.11 |
|  | No | 42,432 | 21.40 | 44,424 | 21.59 | 12,313 | 6.21 | 14,690 | 7.14 |
|  | *p-value* | <.0001 |  | <.0001 |  | <.0001 |  | <.0001 |  |
| Physical activity | Yes | 18,727 | 20.88 | 19,411 | 20.70 | 4,914 | 5.48 | 5,947 | 6.34 |
|  | No | 30,592 | 22.90 | 30,758 | 23.27 | 8,817 | 6.60 | 10,182 | 7.70 |
|  | *p-value* | <.0001 |  | <.0001 |  | <.0001 |  | <.0001 |  |

<sup>a</sup> Results of the Rao-Scott chi-square test

Table 3 presents the influence of the two types of income levels (Korean and community income levels) on perceived stress and experiences of depression by gender. Both Korean and community income levels were significant factors for perceived stress and depression after adjusting for the impacts of other factors in both 2019 and 2021. The odds ratio (OR) of Korean income level of perceived stress was 1.30 (95% Confidence Interval (CI) 1.30–1.30, p<0.0001) and 1.29 (95% CI 1.28–1.29, p<0.0001) in 2019 and 2021, respectively. According to the community income level, the perceived stress for the low-income level group was 1.26 (95% CI 1.26–1.26) and 1.31 (95% CI 1.31–1.32) in 2019 and 2021, respectively, compared to the high-income level group. The experience of depression was significantly lower in the high-income group than in the low-income group (p<0.0001). According to the community income level, perceived stress and experience of depression were both high in the low-income group (2019 OR 1.55, 95% CI 1.55–1.56, 2021 OR 1.63, 95% CI 1.62–1.63). Comparing before and after the COVID-19 pandemic, perceived stress by income level decreased and increased at the Korean and community income level, respectively. During the COVID-19 pandemic, the magnitude of inequality in the experience of depression increased for both types.

**Table 3.**
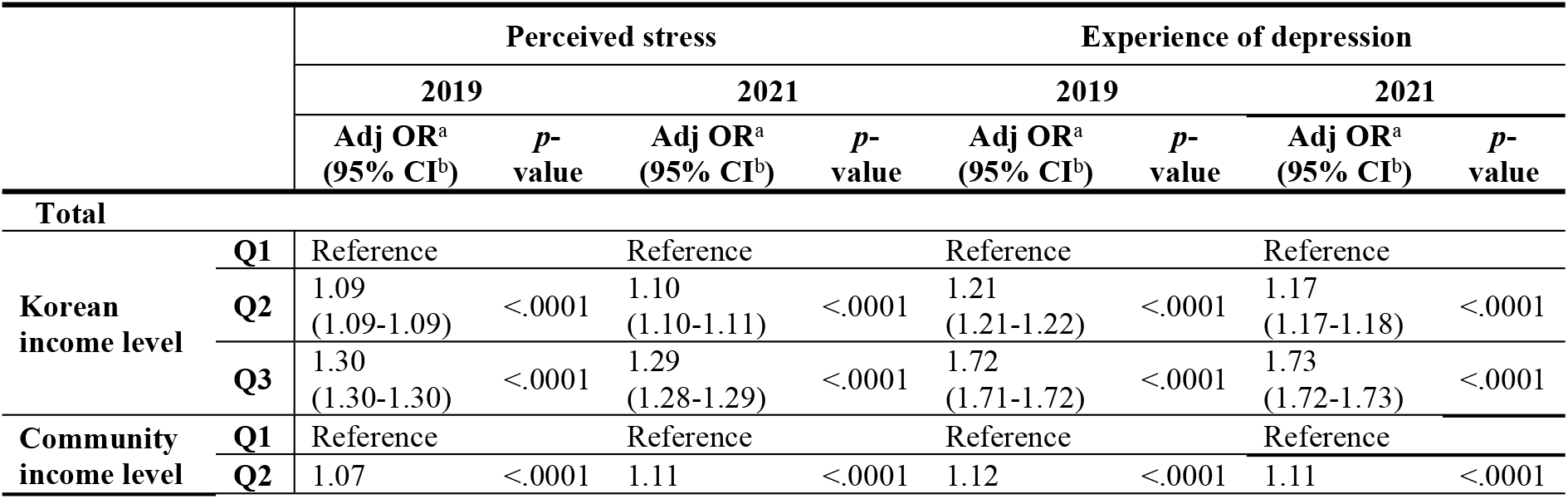
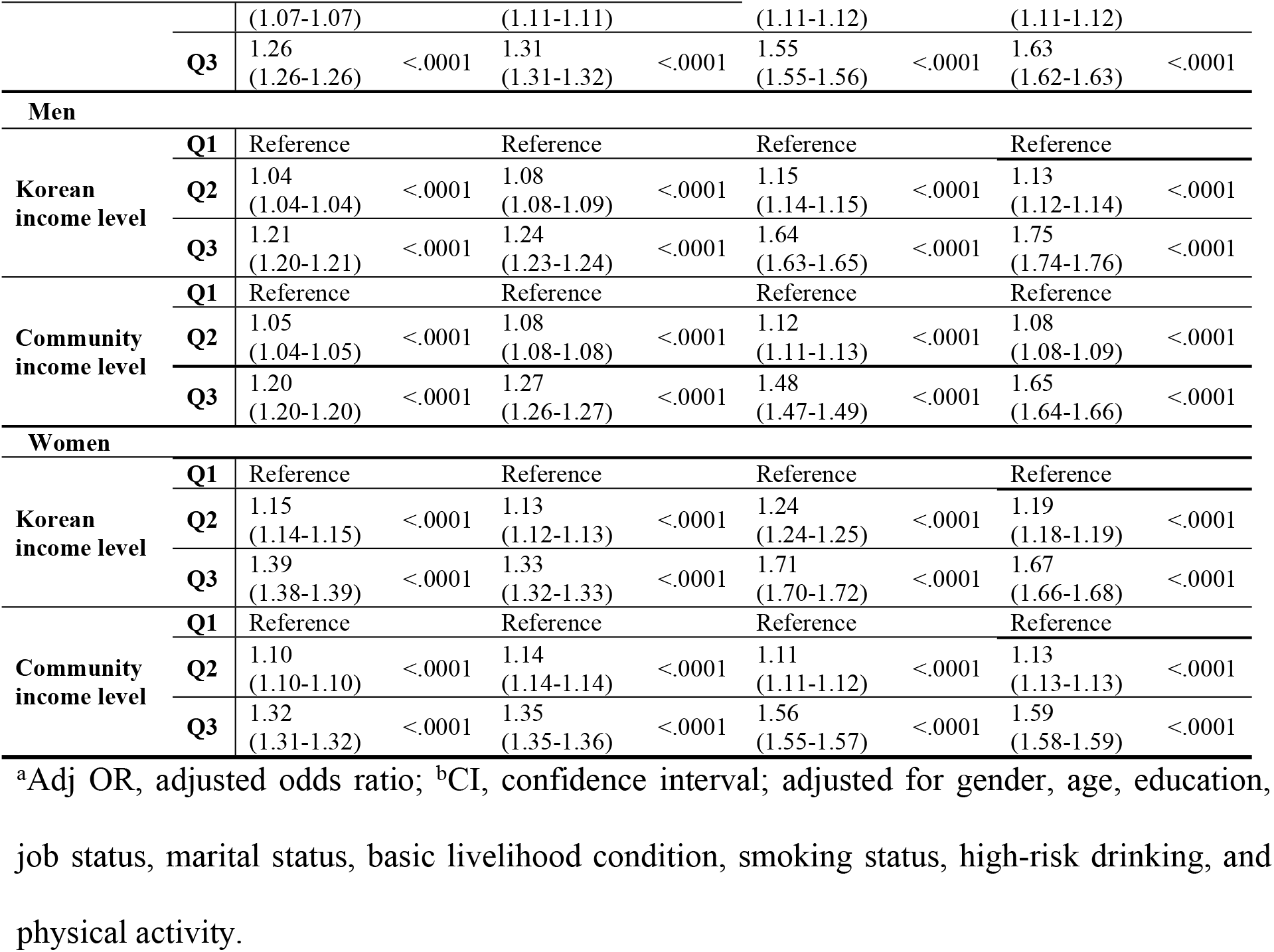
Influence of the two types of income levels (Korean or community income levels) on perceived stress and experience of depression by gender based on complex-sample multivariate logistic regression analysis.

Table 4 shows the results of the subgroup analysis that investigated the income level between the stress perception and experience of depression rates by gender and area of residence. Excluding the Korean income of men living in rural areas in 2021, the magnitude of inequality in stress perception was statistically significant (p<0.0001). The size of the inequality in experiences of depression was statistically significant in both 2019 and 2021, except for men living in rural areas with a Korean income level (p<0.0001). In both 2019 and 2021, the stress perception rate based on Korean income level showed greater inequality in urban areas. Furthermore, community income levels showed greater inequality in urban and rural areas in 2019 and 2021, respectively. The magnitude of inequality in perceived stress by gender was higher for women in both 2019 and 2021. In addition, the magnitude of inequality in the depression recognition was larger for women than for men; however, in 2021, only the magnitude of inequality for men living in rural areas was higher than that for women.

**Table 4.**
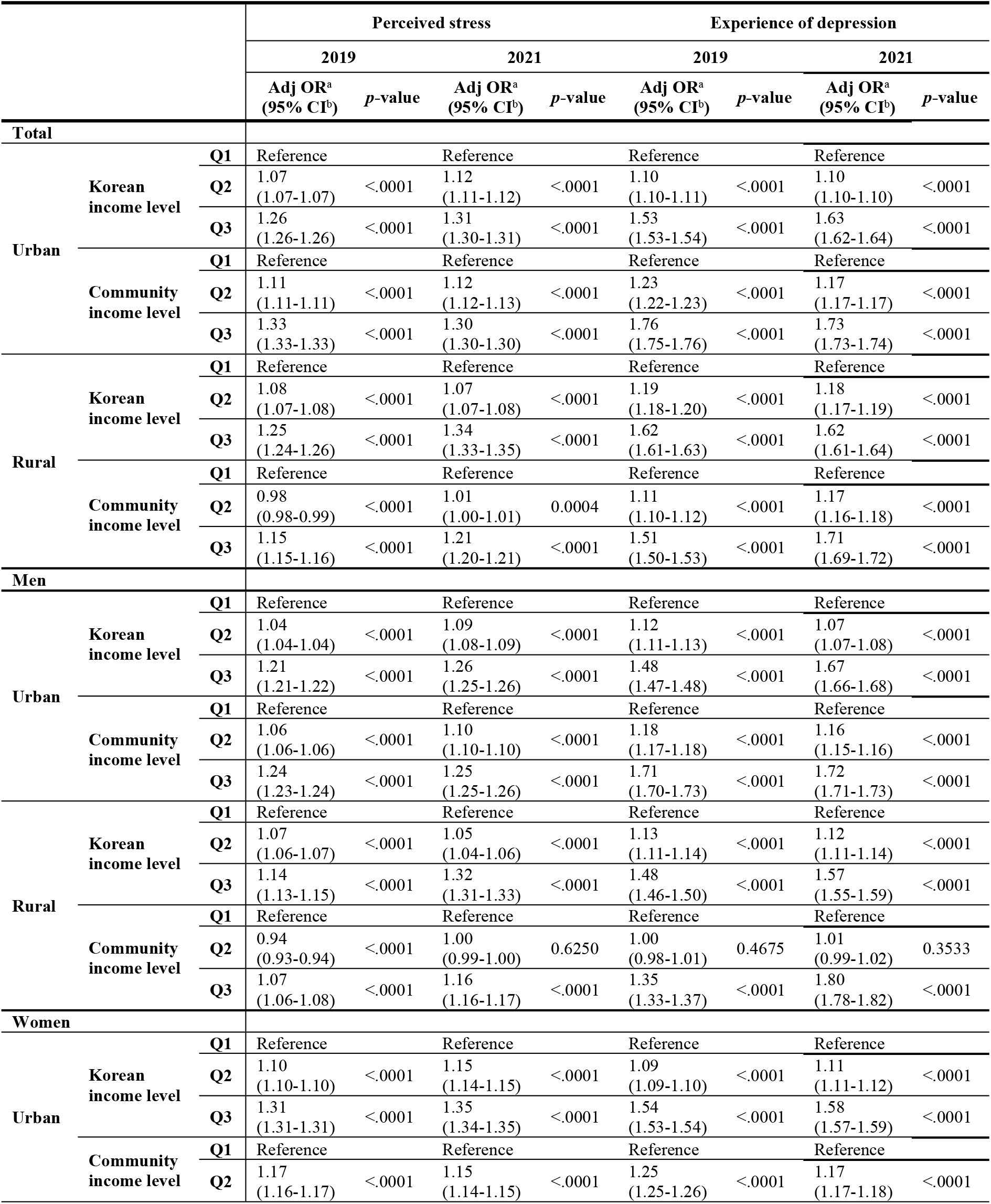
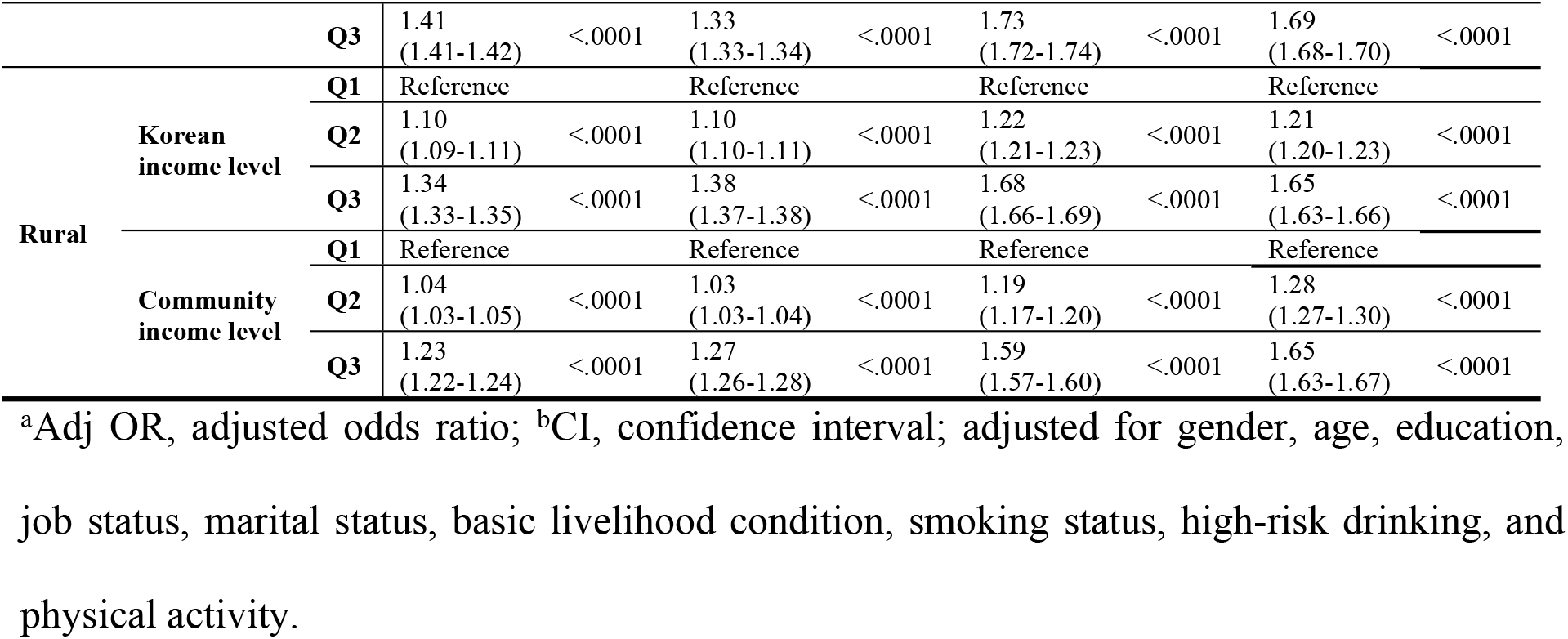
Influence of the two types of income level (Korean or community income level) on perceived stress and experience of depression by gender and region of residence based on complex-sample multivariate logistic regression analysis.

## Discussion

This is the first study in Korea to compare mental health inequality according to relative income at the national and community levels using the CHS during the COVID-19 period. This study intended to provide basic data necessary for policy development to resolve mental health inequality caused by income gaps in the event of large-scale infectious diseases.

During the COVID-19 pandemic, although depression crude rates increased, perceived stress crude rates remained similar. In addition, regarding mental health inequality according to income level, even after adjusting for each independent variable, perceived stress and the experience of depression increased as income level decreased. Hall et al. [19] found that low-income groups had less access to resources for responding to COVID-19 and suffered more economic stress than high-income groups. These economic difficulties reportedly have a negative impact on daily life and mental health [19,40,41]. As a result of measuring perceived stress as income inequality at the national level, it decreased after the COVID-19 outbreak, while income inequality at the community level increased after the outbreak of COVID-19. When measuring inequality by income level, few previous studies have identified mental health inequality at the community income level. Moreover, in some cases, inequality due to the income gap at the community level may be more affected than the income gap at the national level. Aneshensel CS and Sucoff CA [42] and Mair C et al. [43] found that the economic situation (income level), population composition, and characteristics of the residential environment in the area of residence have an effect on depression symptoms. This result is consistent with previous studies that used the relative income gap at the community level as a measure of health inequality and as a significant indicator for identifying the relationship between income and depressive symptoms [15,16].

In this study, perceived stress was more vulnerable in the low-income group compared to the high-income group for both income levels. Experience of depression showed the same results, While women and men in 2019 and 2021, respectively, were susceptible to experiencing depression. The result of women mental health inequality observed in this study was higher than that of men, consistent with the results observed by Almeida et al. [44]. However, after the COVID-19 pandemic, the findings that men experienced higher levels of depression than women were contradictory.

Additionally, this study confirmed the regional characteristics (urban and rural areas) related to changes in mental health inequality according to relative income levels during the COVID-19 pandemic. The effect of relative income level on perceived stress rate was found to be more pronounced in urban areas than in rural areas. On the contrary, the effect of relative income level on the depression rate was found to be weaker and more vulnerable in urban and rural areas, in terms of Korean and community income levels, respectively. This lack of consistent results has been reported in previous studies, as the impact on mental health inequality in urban and rural areas is conflicting [45–49]. In urban areas, social distancing is strongly practiced because of the high number of infected people, which may increase mental health inequality. Furthermore, mental health may be more vulnerable in rural areas because of the lack of information, access, and social support. These results indicate that the relative income level during the COVID-19 pandemic can be expected to have different effects on mental health depending on regional characteristics; however, additional research needs to be conducted in the future.

The limitations of this study are as follows. First, the perceived stress and experience of depression used as outcome variables are values that record responses in the form of self­report rather than medical diagnosis, which raises the possibility of bias. However, because items most frequently used in national surveys were used, the possibility of comparison with other studies related to this topic is high. Second, changes in direct mental health inequalities across the subperiods of the COVID-19 pandemic were not identified. In Korea, the COVID-19 pandemic has usually been divided into three periods: the initial epidemic, delta mutation, and omicron epidemic phase. It is necessary to identify the difference in mental health inequality according to the income gap within the detailed epidemic period because the response strategies differ depending on the size of the epidemic and the impact of restrictions on socioeconomic activities, such as social distancing. In addition, future studies will need to address how inequality will change even after the end of COVID-19.

## Conclusions

This study identified aspects of mental health inequality according to relative income during the COVID-19 pandemic and changes in inequality according to two types of income levels. This study indicated inequality in mental health according to income level and differences based on gender and residential area. To alleviate mental health inequality, income inequality should be improved, and mental health policies should be intensively implemented, especially for socioeconomically unequal population groups.

## Data Availability

https://chs.kdca.go.kr

## Acknowledgments

This study was supported by a 2022 research grant from Pusan National University Yangsan Hospital.

